# SurvMaximin: Robust Federated Approach to Transporting Survival Risk Prediction Models

**DOI:** 10.1101/2022.02.03.22270410

**Authors:** Xuan Wang, Harrison G Zhang, Xin Xiong, Chuan Hong, Griffin M Weber, Gabriel A Brat, Clara-Lea Bonzel, Yuan Luo, Rui Duan, Nathan P Palmer, Meghan R Hutch, Alba Gutiérrez-Sacristán, Riccardo Bellazzi, Luca Chiovato, Kelly Cho, Arianna Dagliati, Hossein Estiri, Noelia García-Barrio, Romain Griffier, David A Hanauer, Yuk-Lam Ho, John H Holmes, Mark S Keller, Jeffrey G Klann, Sehi L’Yi, Sara Lozano-Zahonero, Sarah E Maidlow, Adeline Makoudjou, Alberto Malovini, Bertrand Moal, Jason H Moore, Michele Morris, Danielle L Mowery, Shawn N Murphy, Antoine Neuraz, Kee Yuan Ngiam, Gilbert S Omenn, Lav P Patel, Miguel Pedrera-Jiménez, Andrea Prunotto, Malarkodi Jebathilagam Samayamuthu, Fernando J Sanz Vidorreta, Emily R Schriver, Petra Schubert, Pablo Serrano-Balazote, Andrew M South, Amelia LM Tan, Byorn W.L. Tan, Valentina Tibollo, Patric Tippmann, Shyam Visweswaran, Zongqi Xia, William Yuan, Daniela Zöller, Isaac S Kohane, The Consortium for Clinical Characterization of COVID-19 by EHR (4CE), Paul Avillach, Zijian Guo, Tianxi Cai

## Abstract

**Objective:** For multi-center heterogeneous Real-World Data (RWD) with time-to-event outcomes and high-dimensional features, we propose the SurvMaximin algorithm to estimate Cox model feature coefficients for a target population by borrowing summary information from a set of health care centers without sharing patient-level information.

**Materials and Methods:** For each of the centers from which we want to borrow information to improve the prediction performance for the target population, a penalized Cox model is fitted to estimate feature coefficients for the center. Using estimated feature coefficients and the covariance matrix of the target population, we then obtain a SurvMaximin estimated set of feature coefficients for the target population. The target population can be an entire cohort comprised of all centers, corresponding to federated learning, or can be a single center, corresponding to transfer learning.

**Results:** Simulation studies and a real-world international electronic health records application study, with 15 participating health care centers across three countries (France, Germany, and the U.S.), show that the proposed SurvMaximin algorithm achieves comparable or higher accuracy compared with the estimator using only the information of the target site and other existing methods. The SurvMaximin estimator is robust to variations in sample sizes and estimated feature coefficients between centers, which amounts to significantly improved estimates for target sites with fewer observations.

**Conclusions:** The SurvMaximin method is well suited for both federated and transfer learning in the high-dimensional survival analysis setting. SurvMaximin only requires a one-time summary information exchange from participating centers. Estimated regression vectors can be very heterogeneous. SurvMaximin provides robust Cox feature coefficient estimates without outcome information in the target population and is privacy-preserving.

## 1 INTRODUCTION

Electronic health records (EHR) have been widely adopted in the U.S. and other countries [1-5]. The EHR contains a wealth of patient medical information collected over time by health care providers, and common structured data types include demographics, diagnoses, laboratory test results, medications, and vital signs. Given its longitudinal nature, EHR data have been utilized for various research purposes, including survival analysis [6-8]. For example, the Cox proportional hazards model is used commonly and has been applied to EHR risk prediction [9].

With the increasing availability of EHR data, there is a great interest in integrating knowledge from a diverse range of health care centers to improve generalizability and accelerate discoveries. There now exist multiple collaborative consortia each composed of diverse health care centers seeking to leverage their EHR data in unison. For example, the Consortium for Clinical Characterization of COVID-19 by EHR (4CE consortium) is an international research collaborative that collects patient-level EHR data to study the epidemiology and clinical course of COVID-19 [10]. The consortium comprises more than 300 hospitals across seven countries with 83,178 patients, representing a broad range of multi-national health care centers serving diverse patient populations.

However, EHR data obtained from multiple diverse health care centers often exhibit a high degree of heterogeneity due to variability in EHR and data warehouse platforms, patient populations, health care practices, coding, and documentation. Further, patient-level data often cannot be shared directly between health care centers in a timely manner due to patient and institutional privacy laws [11]. Thus, there is a need for robust analytic strategies to overcome the barriers to conduct multi-center EHR studies.

Our objective is to jointly leverage multi-center, high-dimensional EHR data to make more precise inferences for a target population in the survival analysis setting by sharing only summary statistics obtained from each center, such as Cox feature coefficients and covariance matrices. The target population may be the entire population inclusive of all centers, a subset of centers, or a new, separate population. Integrative analysis approaches that only require individual sites to share summary statistics are often referred to as federated learning [12-14].

Most existing federated learning methods focus on settings with a small number of predictors and/or homogeneous settings where the underlying predictive models are shared across sites [12-16]. In addition, existing methods generally require several rounds of communication between sites, which can be inefficient and labor-intensive. To ensure transportability of models across sites, transfer learning methods have been proposed to transfer knowledge from separate but related centers to provide robust and precise estimates for patients in a new center. This approach has widespread applications in medical studies such as drug sensitivity prediction, integrative analysis of “multi-omics” data, and natural language processing [17-20]. However, most transfer learning methods require outcome labels from the target population, which may be difficult and expensive to obtain, and do not consider the federated learning scenario where individual-level data cannot be shared across sites. In the absence of outcome labels in the target population, transfer learning methods require stringent assumptions that the target and source populations share the same underlying risk model, leading to potential transfer failure when the risk model for the target population is similar to only a subset of source populations [21].

With heterogeneous training datasets from multiple centers, one potential limitation of the existing federated transfer learning methods is that the performance of the prediction model can vary substantially across centers. Thus, although the overall performance may be satisfactory, the performance of the model in a particular center might be low. Moreover, when trained models are applied to a new population, transferability and portability are not guaranteed. To improve the robustness of prediction models, the maximin effect approach was first proposed in [11,14,15], and used as a metric to build a robust prediction model for continuous outcomes across heterogeneous training datasets [22-24]. Instead of optimizing the average performance across all training datasets, the maximin effect method aims to train a model that maximizes the minimum gain over the null model among all training datasets. The maximin approach was further extended to a setting that allows for covariate shift between the source and target populations [25]. The group distributional robustness optimization in [20] is closely related to the maximin effect, which builds a robust prediction model by minimizing the worst-case training loss over a class of distributions [26]. The maximin projection has been developed in [21] to construct the optimal treatment regimen for new patients by leveraging training data from different groups with heterogeneity in optimal treatment decision [27].

In this paper, motivated by the maximin algorithm for continuous outcomes in [11,27], we propose a maximin transfer learning algorithm for predicting a survival outcome (SurvMaximin) in a target population with high-dimensional features by robustly combining multiple prediction models trained in different source populations [22-25]. This algorithm only requires sharing of summary statistics across centers and can easily accommodate high-dimensional features. SurvMaximin can be viewed as a robust federated approach to transfer models trained at multiple external centers to a target population, so we refer to it as a federated transfer learning method. SurvMaximin differs from existing transfer learning methods in that it does not require the target population to share the same underlying model with the source population, a highly desirable property when learning with multiple heterogeneous health care systems. The training of the SurvMaximin algorithm also does not require the target population to have gold-standard outcome labels.

## 2 METHODS

### 2.1 SurvMaximin: Federated Robust Transfer Learning for Survival Outcomes

The main aim of the SurvMaximin algorithm is to derive a robust risk prediction model for an unlabeled target population based on labeled data from the *L* source populations under the data sharing constraints. Suppose there are *L* source populations, indexed by *l* ∈ {1,…,*L*}, representing *L* studies and one target population, denoted by *Q*. The observed data from the *l*th source population consists of *n*_*l*_ independent and identically federated random vectors 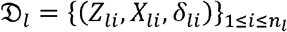, where *Z*_*li*_ denotes the *p*-dimensional standardized baseline risk factors with *p* potentially large relative to the sample sizes, the censored survival times are observed as *X*_*li*_ = (*T*_*li*_, *C*_*li*_) and *δ*_*li*_ = *I*(*T*_*li*_ ≤*C*_*li*_) with *T*_*li*_ and *C*_*li*_ denoting the survival time and follow up time for the *i*th subject in the lth population, respectively. In the target population *Q*, only the baseline features 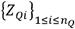 are observed. We assume that the survival time in the *l*th source population, *T*_*l*_, given the baseline features *Z*_*l*_ follows a Cox proportional hazards model, 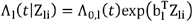, which can be equivalently expressed as

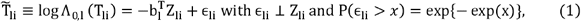

where *Λ*_*l*_ (*Z*_*li*_) is the conditional cumulative hazard function given Z for the *l*th population, *Λ*_0,*l*_(*t*) is the cumulative baseline hazard function, 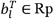 denotes the vector of unknown log hazard ratio parameters associated with the risk factors *Z*. We assume that distributions of the baseline risk factors, hazard ratio parameters, and the baseline hazard functions may vary across the source populations due study heterogeneity. Similarly, we assume that the survival time from the target population, *T*_*Q*_, follows a Cox model with unknown baseline cumulative hazard *Λ*_0, *Q*_ (·) and feature effect function *β*_*Q*_ :

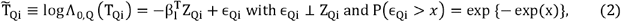

The SurvMaximin algorithm aims to identify a robust approximation to *β*_*Q*_ based on the estimated hazard ratio parameters trained from {𝔇_*l*_}_1≤*l≤L*_ as well as the target feature distribution. Due to the lack of gold standard labels on *Q* and the unspecified heterogeneity among {*b*_*l*_}_1≤*l*≤*L*_ and *β*_*Q*_, the target *β*_*Q*_ cannot be identified with the observed data. Instead of targeting *β*_*Q*_ directly, the central idea of the SurvMaximin algorithm is to identify an approximation to *β*_*Q*_ that maximizes the minimum reward across all *L* source populations. Following [27], we define the hypothetical outcomes for the *n*_*Q*_ subjects in Q generated from the lth source models as

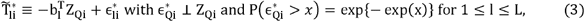

where 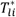 can be viewed as the hypothetical outcome (transformed survival time) if the individual *Z*_*Qi*_ was assigned to the *l*th source population. Then we define a robust prediction model as

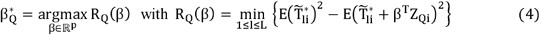

the expectation is taken with respect to *Z*_*Qi*_ and 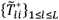 defined in (3). Such a covariate shift maximin effect was defined in [27] for the linear model and is now extended to the Cox regression model. We shall note that 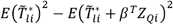 is a reward function of *β*, which represents the variance of 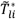 explained by the linear prediction −*β*^*T*^*Z*_*Qi*_. Our targeted maximin effect is maximizing the adversarial reward *R*_*Q*_(*β*) across the *L* groups. The SurvMaximin estimate 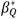 leads to a robust prediction model since the optimization in (4) guards against the worst case scenario. The maximin effect can be interpreted from an adversarial perspective [14]: in a two-side game, we select an effect vector *β* and the counter agent then chooses the most challenging scenario for this *β*; that is, choose the source population such that *β* has the worst predictive performance. Our goal is to choose *β* such that the worst case reward with respect to predicting the transformed survival time returned by the counter agent is maximized.

It is not difficult to see that *R*_*Q*_ (*β*) can be equivalently expressed as:

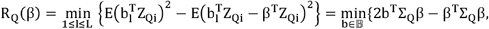

where Σ_Q_ = E[Z_Qi_(Z_Qi_)^T^], and 𝔹 = {b_l_,l = 1,…,L}. Following [27], we may show that the maximin effect 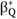 as defined by (4) can be expressed as a weighted average of {b_l_}_1 ≤ l ≤ L_,

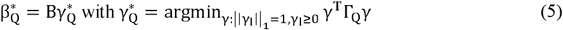

where B_p ×L_ = [b_1_,…,b_L_], ‖ · ‖ _q_ denotes the L_q_ norm, and the minimization above is restricted to the simplex in L-dimension space. The optimal aggregation weight 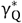 in (5) depends on both {b_l_}_1 ≤ l ≤ L_ and the covariance matrix Σ_Q_ for the target population. The identification equation (5) reveals an important geometric interpretation of the maximin effect: 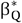 is the point that has the smallest distance to the origin and lies on the convex combination of the regression vectors {b_l_}_1 ≤ l ≤ L_ [22]. The maximin estimator tends to shrink the components of {*b*_*l*_}_1 ≤ *l* ≤ *L*_ whose estimated coefficients vary with different signs across studies to zero and is not as sensitive to the inclusion of sites with an extreme hazard ratio regression vector [22]. In the transfer learning setting, we incorporate the target distribution Z_Q_ into the definition of the distance.

### 2.2 Implementation of the SurvMaximin Algorithm

The SurvMaximin algorithm involves three key steps: (I) locally train the prediction model for each of the *L* source sites to obtain {**b**_*l*_}_1 ≤ *l* ≤ *L*_; (II) estimate the covariance matrix **Σ**_*Q*_ and obtain a similarity matrix among 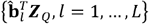, denoted by **Γ**_*Q*_; and (III) obtain the final SurvMaximin estimator as an optimal linear combination of 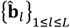 according to (5). The schema of the SurvMaximin algorithm is shown in Figure 1.

**Figure 1:**
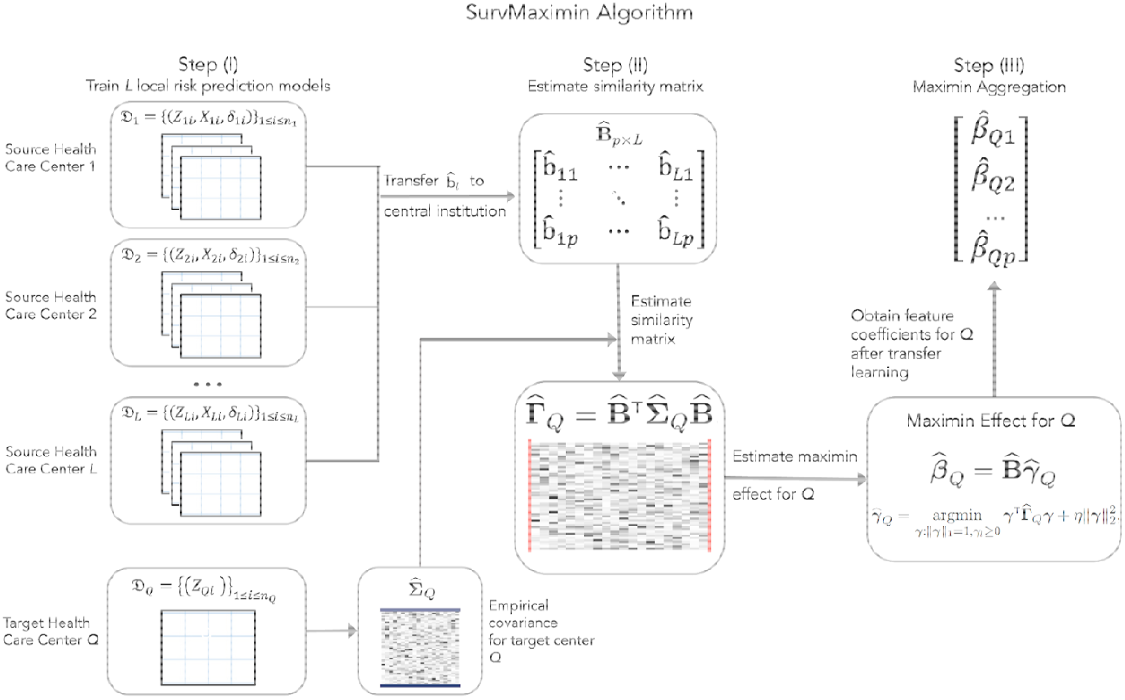
Schematic of SurvMaximin algorithm for federated transfer learning.

#### Step I: Training L local risk prediction models

We first obtain as the maximizer of the penalized partial likelihood where is the log partial likelihood associated with and is the elastic net penalty function, which is frequently used to overcome high dimensionality and collinearity of features with α = 1 corresponding to the standard LASSO and α = 0 corresponding to the ridge penalty [28]. The non-negative penalty parameter can be selected via standard tuning criteria including the AIC, BIC or cross-validation.

#### Step II: Estimate the similarity matrix among 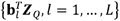

We estimate the similarity matrix of 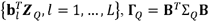, as 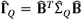, where 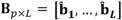 and 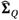 is the empirical variance covariance matrix of ***Z***_*Q*_ estimated based on the unlabeled target population data.

#### Step III: Maximin aggregation via (5)

Finally, we obtain the SurvMaximin aggregated log hazard ratio estimator as

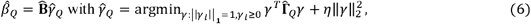

where η ≥ 0 is the tuning parameter and the ridge penalty is included to account for the potential high collinearity among 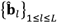. See Supplementary Materials for additional information on data adaptive approach to selecting η. In practice, we find that when there is some heterogeneity observed as in our 4CE studies, setting η = 0 works well and the results are not sensitive to the choice of η when a relatively small η is chosen.

### 2.3 Transfer to a Target Site with Missing features

A substantial challenge in transfer learning across different health care centers is that certain risk predictors, such as laboratory test results or demographic information, may be available in one center but not in a different center. For example, in the 4CE Consortium, all U.S. centers report data on race while European centers do not, causing race data to be entirely missing for European centers. To transport a risk prediction model for a target center Q with only a subset of features available, one may fit a reduced model limited to only the available features for each source center and transport the reduced risk models from the source centers. When the target center changes, essentially we will need to retrain the model at each source center according to the feature availability of the target center. Such an approach is not computationally efficient as each center needs to fit multiple models, and also increases the number of communications required across centers.

To enable transfer learning in the context of differential feature availability, we propose a simple projection approach that only requires each source center to additionally compute the empirical covariance matrix of the features, 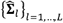, where 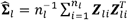. Let 𝒜 ⊂ {1,…,*p*} index features that are available at the target site, 𝒜^*c*^ = {1,…,*p*}\ 𝒜. Let ***Z***^[𝒜]^ denote the subvector ***Z*** corresponding to 𝒜. The key step is to project 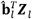 to the subspace spanned by 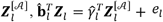, and and predict *T*_*l*_ based on 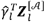. Since the features are all assumed to be centered, we obtain 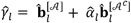 and 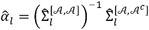, where 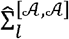 and 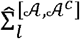 denote the submatrices of Σ_*l*_ corresponding to { 𝒜, 𝒜 } and { 𝒜, 𝒜^*c*^}. The final SurvMaximin estimator for the feature effects of 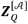 can be constructed by replacing 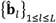 with 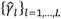 and 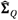 with 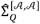.

### 2.4 Validation of SurvMaximin Algorithm

We validated the performance of SurvMaximin in federated transfer learning using both simulation studies and a real-world study where we transported COVID-19 mortality risk prediction models to target centers using EHR data from hospitalized patients with COVID-19.

#### 2.4.1 Simulation Studies

Simulation studies were conducted to assess the performance of SurvMaximin and to compare its performance against existing federated learning methods. Since SurvMaximin transports a risk prediction model to a future target center without survival outcomes, use other federating learning methods that also do not require supervised training on the target data as comparisons. Specifically, we consider the standard random effect meta-analysis estimator (herein referred to as Meta); the One-shot Distributed Algorithm (ODAC) for the Cox model [26]; and the locally trained risk prediction model with varying training sizes of *n*_*Q*_ = 200, 400, and 600. We considered simulation scenarios with *L* = 15 centers each with sample size 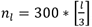, and *p* = 20 or 50 features in the risk prediction model.

We generated ***Z***_*l*_ from a multivariate normal distribution MVN (0,Σ), where Σ is either 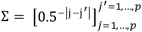 with an autoregressive correlation (AR) structure, or where Σ is a compound symmetry covariance matrix with variance 1 and covariance 0.5. We then generated ***Z***_*Q*_ from MVN (0,Σ_Q_) with Σ_*Q*_ = 0.1 + Σ. Subsequently, we generated *T*_*l*_ and *T*_*Q*_ from:

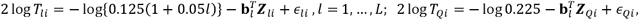

where *ϵ*_*li*_ and *ϵ*_*Qi*_ were generated from extreme value distributions. We let

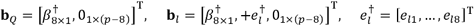

and consider a range of scenarios for *β*^†^ and {*e*_*l*_ }_*l*=1,…,*L*_ to explore how the signal strength, the heterogeneity among the source sites, as well as the degree of similarity between the target site and the source sites affect the performance of SurvMaximin relative to other methods. Specifically, we consider *β*^†^ = [0.5,0.4,0.3,0.2,−0.2, −0.3, −0.4, −0.5]^T^ and [0.25,0.2,0.15,0.1, −0.1, −0.15, −0.2, −0.25]^T^ to represent moderate and weak signals. We consider two settings for 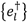 under each with three levels of heterogeneity among source sites for *e*_*l*_. In setting (I), we let *e*_*lj*_ = *τ*{1(*l* ≤ 5) + 3*I*(*l* > 5)}(−1)^*j*^, which results in the first 5 sites being more similar to the target site than the remaining 10 sites. In setting (II), we let *e*_*lj*_ = *τ*(*l* − 1) such that a majority of the source sites are substantially different from the target site. We let τ = 0.05, 0.1, and 0.2 to reflect a low, medium and high degree of heterogeneity among the source sites. As τ increases, the target site also becomes more dissimilar to the source sites. We generated censoring time from Exponential (3) distribution, leading to about 20% to 30% event rates across the *L* source sites.

To evaluate the performance of SurvMaximin for missing features, we considered setting (I) with moderate signal, covariance matrix Σ being AR (1), p = 20, and τ = 0.05, 0.1, 0.2. We let the first feature of the target site being missing and calculate the projected SurvMaximin estimator as described in Section 2.3, denoted by SurvMaximin_project_. For comparison, we also fitted penalized Cox models to each site with covariates ***Z***^[𝒜]^ with 𝒜= {2,…,*p*} to obtain the corresponding effect estimates 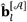 for ***Z***^[𝒜]^; and then constructed SurvMaximin based on 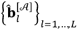. As naïve benchmarks, we additionally constructed the ODAC and Meta models based on ***Z*** and transported these models to the target site by removing the component associated with the first covariate. Such naive approaches are often adopted in practice due to the inability to refit the reduced models on source sites.

We evaluate the overall performance of the estimated risk score from each method in predicting the survival time *T*_*Q*_ for the target site, based on the survival C-statistic with a truncation time close to the largest observed survival time from the target site [29]. We estimate the C-statistics based on an independent validation data of size *N*_*Q*_ = 2,000 generated from the target distributions. For each configuration, we summarize results based on 500 iterations.

#### 2.4.2 Improving Cross-system Portability of COVID-19 Mortality Risk Prediction Models with SurvMaximin

We further validated the performance of SurvMaximin by deriving robust and transportable mortality risk prediction models for patients hospitalized with COVID-19 using international, multi- institutional EHR data from the 4CE consortium [10,16]. Baseline risk factors and mortality information were available for 83,178 patients from *L*_0_ = 15 participating health care centers of the consortium across three countries: France, Germany, and the US. Eligibility criteria for the study included a positive SARS-CoV-2 reverse transcription polymerase chain reaction (PCR) test result; an admission date between March 1, 2020 and January 31, 2021; and the admission occurred 7 days before to 14 days after the date of their first positive PCR test result recorded in their EHR. Each health care center performed analyses locally and then reported summary results to the central institution. We consider each of the individual health care center as a potential target population and sought to derive a mortality risk prediction model that is transportable to this population from multiple external models. Given the multinational nature of our data, we anticipated a significant amount of between health care center heterogeneity in their mortality risk models.

Baseline risk predictors considered include: age groups (18-25, 26-49, 50-69, 70-80, 80+) sex, and race (White, Black, Asian, Hispanic and other); the pre-admission Charlson comorbidity index (CCI) derived from diagnostic codes; and laboratory test values at admission [30]. We focused on ten commonly measured laboratory tests (with missing rates < 30%), including C-reactive protein (CRP), albumin, aspartate aminotransferase (AST), AST to alanine aminotransferase ratio (AST/ALT), total bilirubin, creatinine, D-dimer, white blood cell count (WBC), lymphocyte count, and neutrophil count. Values of AST, D-dimer and CRP were log-transformed due to their skewed distributions. Missing baseline laboratory values and CCI were imputed via the multivariate imputation by chained equation method and averaged over five imputed sets [31]. In total, we considered *p* = 19 potential risk predictors. A few predictors, including race data for the European centers, were not available (Supplementary Figure S3). When a variable is not ascertained at a site, the local Cox model fitting excluded it. We derived and evaluated prediction models for all-cause mortality by 3, 7 and 14 days after the admission date. We excluded patients who died on the day of admission in the survival analysis.

For each *L*_0_ = 15 health care centers, we transported mortality risk prediction models trained from external analyses via SurvMaximin to the patient population in this center. Specifically, for the lth healthcare center, we fit LASSO penalized Cox models to estimate the effect of *Z*_*l*_ coefficients 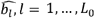 on survival outcome. For *l* = 1,…,*L*_0_, we let the *l*th site be the target site and then train the SurvMaximin algorithm based on the source data from all remaining sites that have predictors 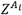 ascertained, where 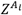 denotes the covariate vector that is available at site *l*. We use the proposed projection method when the target center had an incomplete set of features. After obtaining SurvMaximin risk model for the target center, we compared the SurvMaximin risk model against each of the *L*_0_ supervised locally trained models with respect to the accuracy in predicting *t*_0_ = 3,7,14 day mortality in the target population. We quantify the accuracy for predicting *t*_0_-day mortality based on the area □ under the receiver characteristic curve (AUC). We repeated this analysis for all *L*_0_ = 15 centers each time considering one of them as the target population *Q*.

## 3 Results

### 3.1 Results for simulation studies

Simulation results are summarized in Figure 2 for the moderate signal scenario. In setting (I), where 5 source sites have feature coefficients like the target site, SurvMaximin results in models with accuracy comparable to those from ODAC and Meta when the heterogeneity is low (τ = 0.05, 0.1) and outperforms other methods when the heterogeneity is high (τ = 0.2). Since there are 5 source sites relatively like the target site, the transported model from SurvMaximin attained accuracy higher than the locally trained model with *n*_*Q*_ = 200 and comparable to those trained with *n*_*Q*_ = 600. When *p* or the correlation among the features increases, the estimated models generally attain lower prediction performance. Nevertheless, SurvMaximin remains to attain a robust performance relative to the other federated learning methods across different levels of heterogeneity.

**Figure 2:**
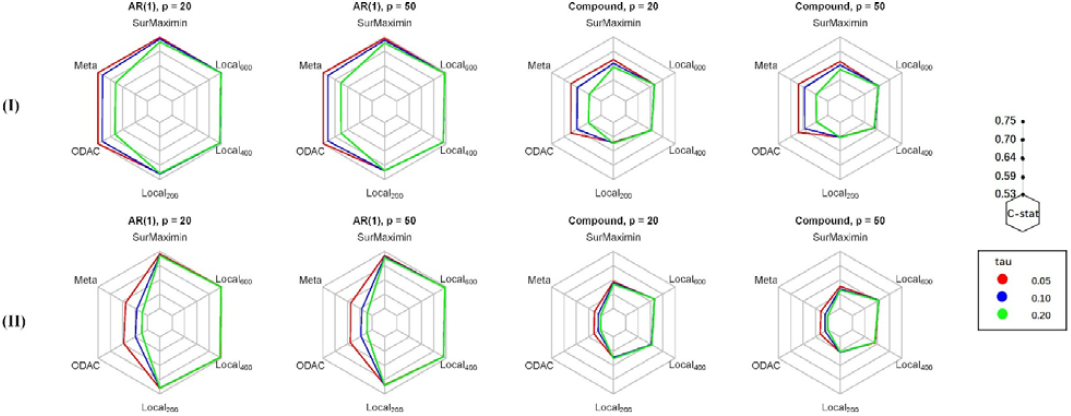
Average C-statistics under settings (I) and (II) with **Σ** being either AR(1) or compound symmetry; *p* = 20 *or* 50; and tau = 0.05, 0.10, or 0.20 (local coefficients heterogenicity) for predicting survival in the target population with risk models trained by SurvMaximin, (Meta), ODAC, as well as supervised penalized Cox regression with *n*_*Q*_ = 200,400,600 labeled target data (Local_200_, Local_400_, Local_600_).

In setting (II), where only 1 other site has similar feature coefficients to the target site, SurvMaximin exhibits a substantially better predictive performance when compared to Meta and ODAC across all settings, further highlighting the robustness of SurvMaximin to varying degrees of similarity between the target site and the source sites. Across all levels of heterogeneity, the Meta and ODAC estimators suffer from very small C-statistics indicating poor predictive performance. We observed similar trends regardless of using *p* = 20, 50 features or covariance matrix structure. Further, the performance of SurvMaximin remains better than the supervised model trained with *n*_*Q*_=200 labeled target site data and comparable to the locally trained models with *n*_*Q*_ = 400 and 600. This suggests that SurvMaximin may improve estimation performance when the target population sample size is small.

With weaker signals, the cross-site heterogeneity is more pronounced, leading to more apparent distinctions between SurvMaximin and other federated methods (Figure S1 of the Supplement). Only when the heterogeneity is very low with τ = 0.05 under Setting (I), all methods perform similarly. Under all other settings, SurvMaximin substantially outperforms ODAC and Meta. With weaker signals, locally trained models also require a larger sample size to attain performances comparable to SurvMaximin. This further illustrates the advantage of transporting existing models in a robust fashion over training a supervised model when the training sample size is not large relative to the feature dimension.

Results for assessing the performance of the projected SurvMaximin algorithm in the presence of missing features are summarized in Figure S2 of the Supplement. The projected SurvMaximin model attains prediction performance comparable to the SurvMaximin model trained by aggregating the locally fit sub-models with Z. Thus, the projection method provides a comparable alternative SurvMaximin estimator when features may be missing for some sites, without the need to unify the set of features for all the centers all the time. The projected SurvMaximin estimator also outperforms the naïve approach of removing the component associated with the first covariate from the ODAC or Meta estimators.

### 3.2 Results for Transporting COVID Mortality Risk Models

For each covariate, we compare the *L*_0_ local estimates of its log hazard ratio to those based on SurvMaximin in Figure 3. While these two sets of estimators are generally consistent, SurvMaximin estimators tend to be more concentrated at the center, while local estimators exhibit higher variability in part due to unstable estimates from some sites. For example, the log hazard ratio (HR) of the age group (18-25) ranges from -6.58 to 0 for the local estimates while the SurvMaximin estimates range from -1.43 to -0.7.

**Figure 3:**
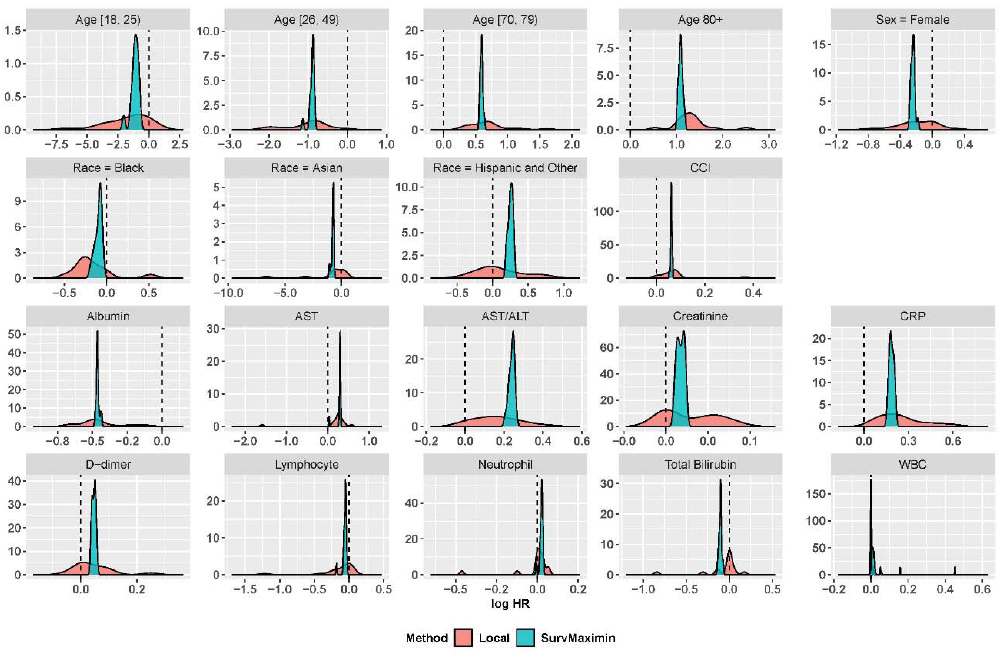
Density plots of two covariate effect estimators for healthcare systems *l* = 1, …, *L*_0_ = 15

The AUC estimates associated with the risk models obtained based on SurvMaximin and local supervised training for predicting 3, 7, and 14-day mortality are shown in Figure 4. For each site, we also compared the AUC of models trained in each of the external sites, the locally trained model, and the SurvMaximin model for predicting 14-day mortality (Figure S4 of the Supplement). The accuracy of risk models transported by SurvMaximin, which does not utilize the outcome information of the target local site, is comparable or even sometimes higher than that of locally trained models. The AUCs of SurvMaximin are more concentrated at a comparatively higher AUC, suggesting the robustness of the SurvMaximin approach.

**Figure 4:**
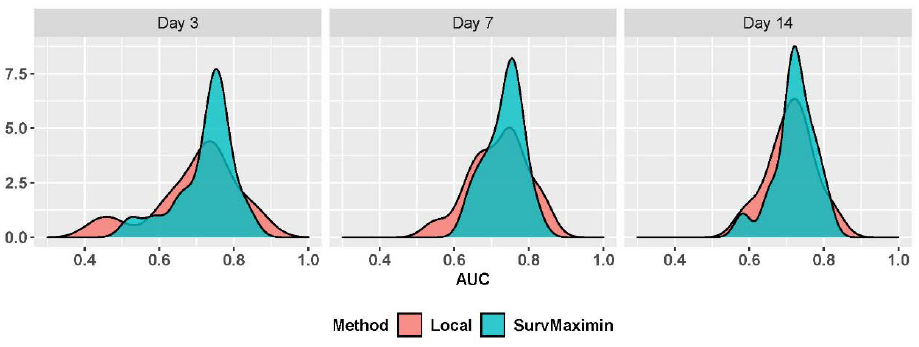
Density plots of two AUC estimators for healthcare systems *l* = 1, …, *L*_0_ = 15.

## 4 Discussion

We proposed the SurvMaximin approach to deriving a robust risk prediction model for a target population by robustly synthesizing information from estimated risk models from multiple sites. For the target site, the SurvMaximin estimator 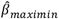 is a linear combination of the coefficient estimators of the local sites 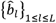, or it lies in the convex hull of all the 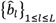 and is closest to a zero point with respect to some distance related to the target population. The method enables us to safely transport a set of existing risk models to a target population in the presence of high cross-site heterogeneity.

Compared with existing federated learning methods, such as Meta and the federated learning methods proposed by [14], the proposed maximin method can handle high-dimensional covariates and is very robust to heterogeneity between sites. It’s also robust to sample size differences and improve the inference when the sample size of the target population is small as seen from the simulation studies. The SurvMaximin algorithm is efficient in time and cost as it only requires one-time sharing of the summary statistics. Compared with existing transfer learning methods, the proposed maximin method can help to preserve the privacy and confidentiality of patients in different centers. Further, it requires less information, only feature information of the target site and feature effect estimators from other sites.

Thus, SurvMaximin is very flexible and generalized such that it can adapt to a variety of scenarios while achieving high accuracy with limited information.

## 5 Conclusion

In this paper, we developed a SurvMaximin covariate effect estimator for multi-center survival data with high-dimensional covariates. Simulation studies and real EHR data analysis show that the proposed estimator achieves high accuracy in a range of settings with different levels of heterogeneity between sites and different sample sizes. The SurvMaximin is a highly flexible and robust approach for multi-center survival analysis, which enables federated learning, transfer learning, as well as federated transfer learning.

## Data Availability

All data produced are...

## Funding

GMW is supported by National Institutes of Health (NIH)/ National Center for Advancing Translational Sciences (NCATS) UL1TR002541, NIH/NCATS UL1TR000005, NIH/National Library of Medicine (NLM) R01LM013345, NIH/ National Human Genome Research Institute (NHGRI) 3U01HG008685-05S2. YL is supported by NIH/NCATS U01TR003528, and NLM 1R01LM013337. KC is supported by VA MVP000 and CIPHER. NGB is supported by PI18/00981, funded by the Carlos III Health Institute. DAH is supported by NCATS UL1TR002240. MSK is supported by NHGRI 5T32HG002295-18. JHM is supported by NLM 010098. MM is supported by NCATS UL1TR001857. DLM is supported by NIH/NCATS CSTA Award #UL1-TR001878. SNM is supported by NCATS 5UL1TR001857-05 and NHGRI 5R01HG009174-04. GSO is supported by NIH U24CA210867 and P30ES017885. LPP is supported by NCATS Clinical and Translational Science Award (CTSA) Award #UL1TR002366. FSJV is supported by NIH/NCATS UL1TR001881. AMS is supported by NIH/ National Heart, Lung, and Blood Institute (NHLBI) K23HL148394 and L40HL148910, and NIH/NCATS UL1TR001420. SV is supported by NCATS UL1TR001857. ZX is supported by National Institute of Neurological Disorders and Stroke (NINDS) R01NS098023. WY is supported by NIH T32HD040128.

## Supplementary Materials

### A. Tuning parameter selection for (6)

For *η* = 0, the optimum reward value is denoted as 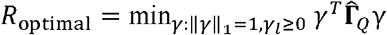. We will choose *η* from an initial range [0, *η*^max^) with

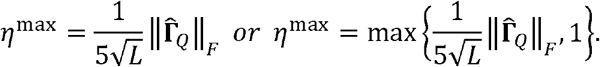

We choose the tuning parameter *η* by checking the minimum eigenvalue of 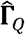, denoted as 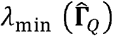. If 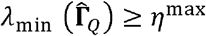, we shall set *η* = 0; otherwise, we choose the largest *η* ∈ (0, *η*^max^) such that the reward of 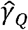 is above 95% of *R*_*optimal*_, where the reward of 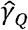 is defined as 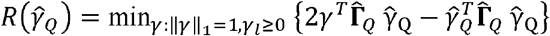; see [27] for further detailed discussion.

**Figure S1:**
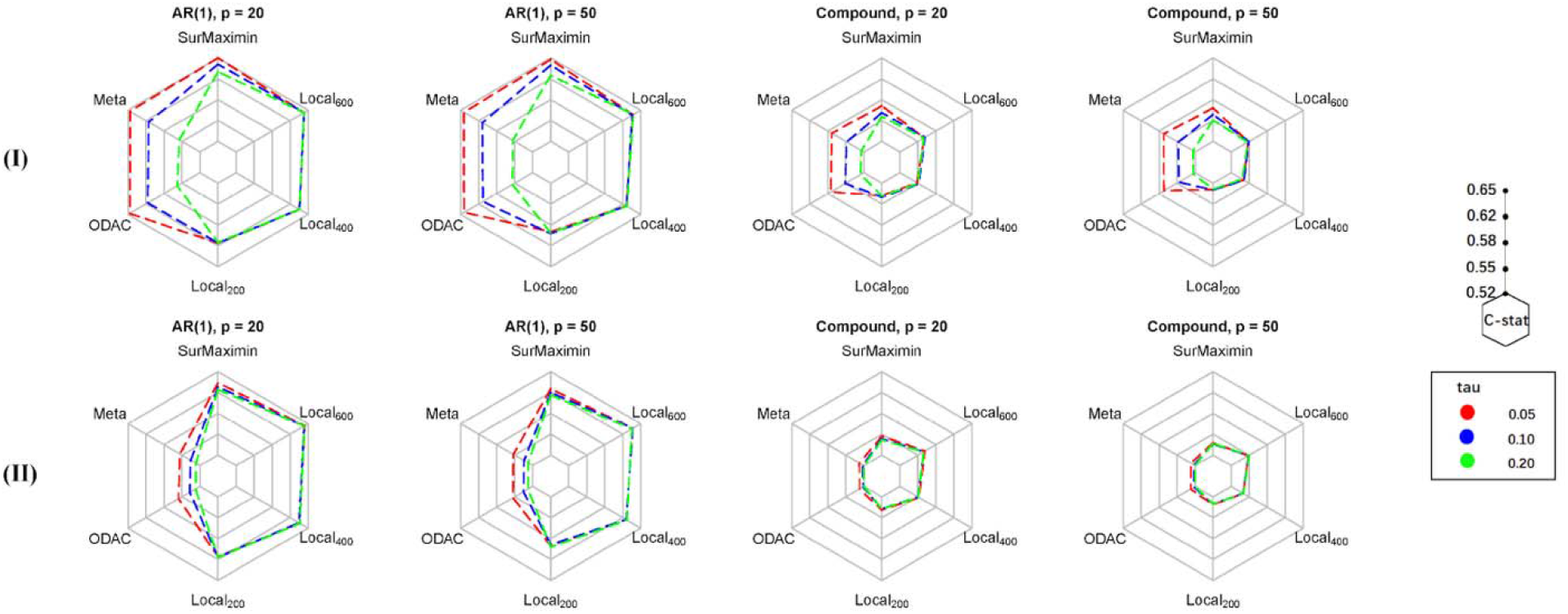
Average C-statistics under settings (I) and (II) with Σ being either AR (1) or compound symmetry, p = 20 or 50, and tau=0.05, 0.1, 0.2 (local coefficients heterogeneity) for predicting survival in the target population with risk models trained by SurvMaximin, Meta, and ODAC, as well as supervised penalized Cox regression with **n**_*Q*_ =200, 400, 600 labeled target data (Local_200_, Local_400_, Local_600_).

**Figure S2:**
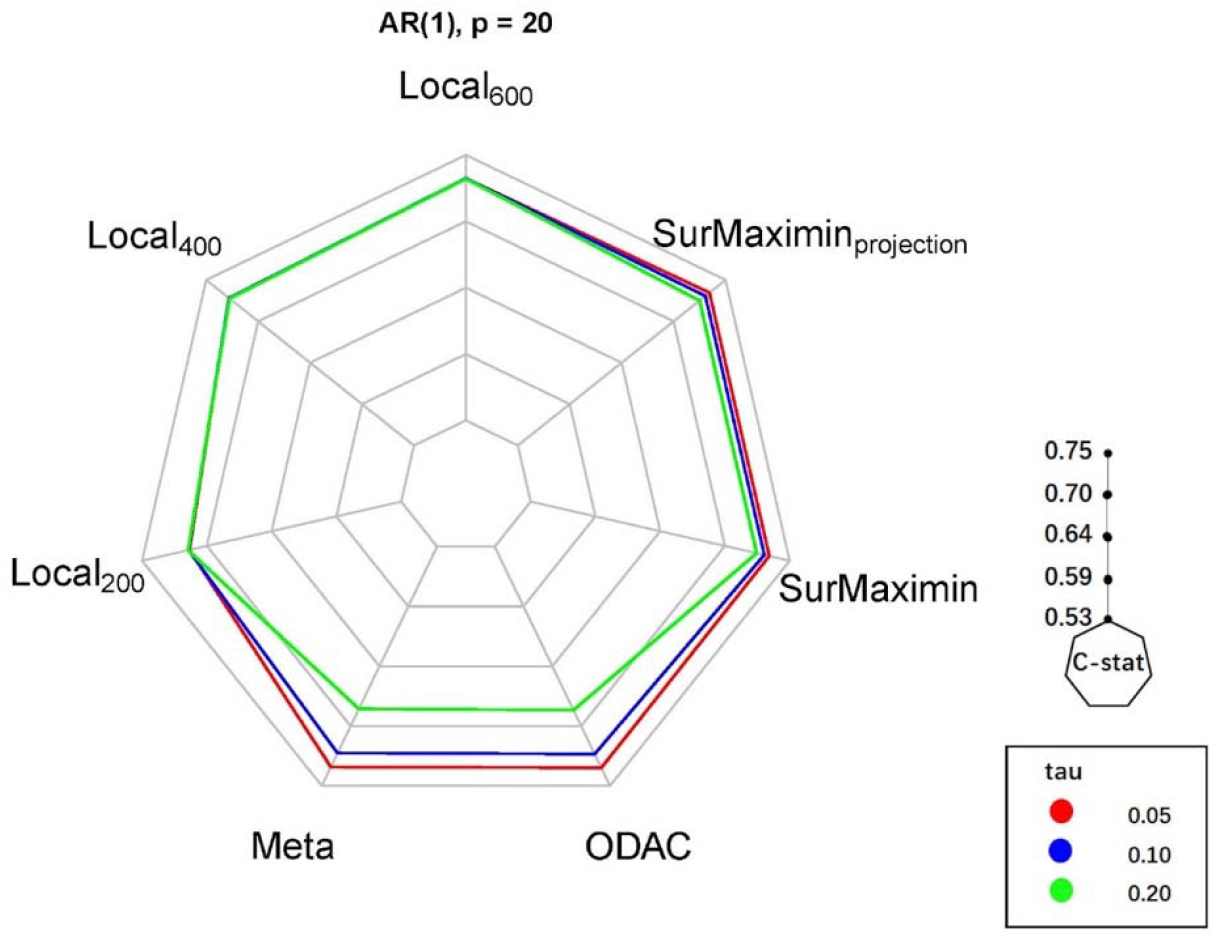
Average C-statistics under settings (I) with Σ being AR (1), p = 20, and tau=0.05, 0.1, 0.2 (local coefficients heterogeneity) for predicting survival in the target population with one missing feature based on risk models trained by SurvMaximin, SurvMaximinproject, Meta, and ODAC, as well as supervised penalized Cox regression with **n**_*Q*_ = 200, 400, 600 labeled target data (Local_200_, Local_400_, Local_600_).

**Figure S3:**
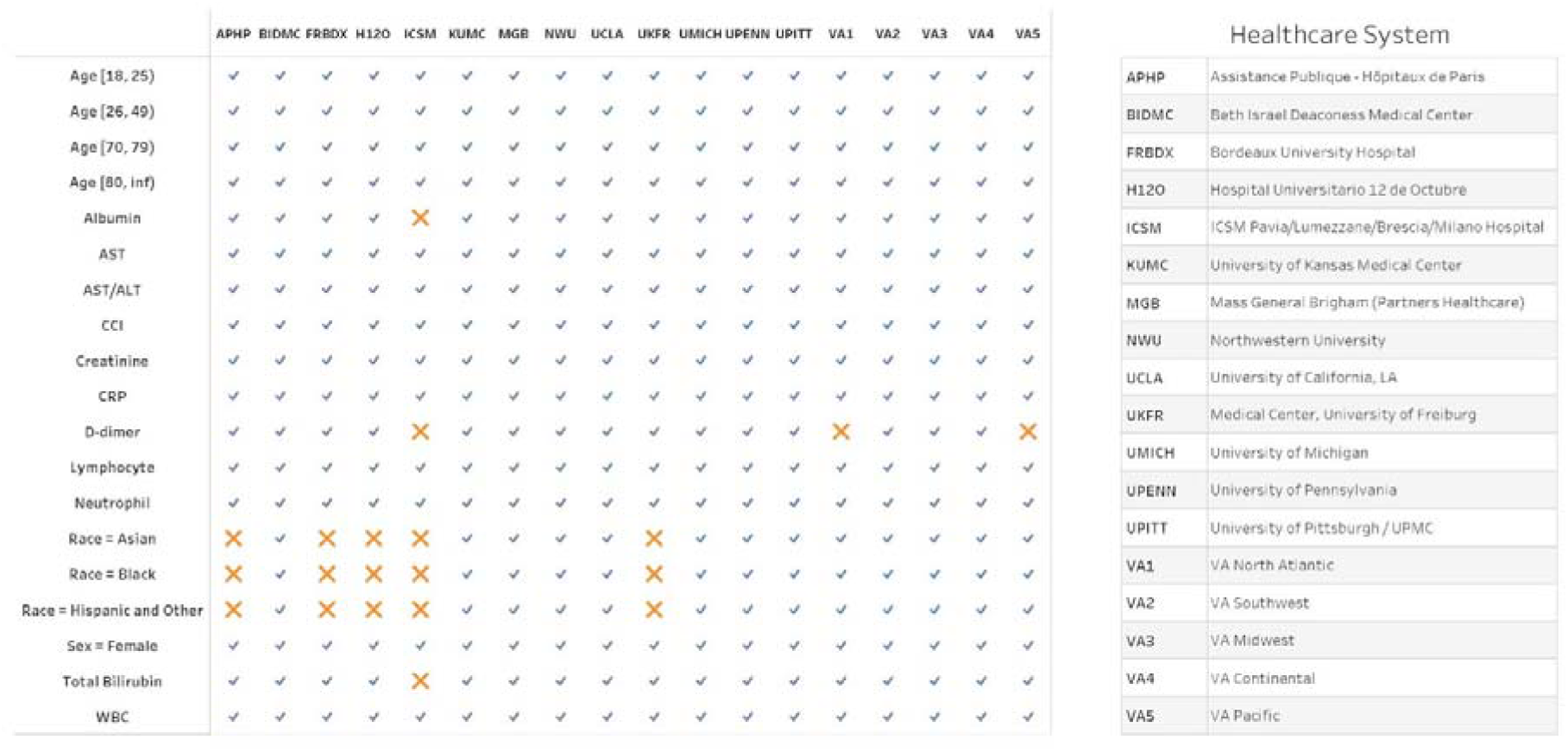
Missing predictors in each site for the COVID-19 Mortality Risk Modeling with 4CE consortium EHR data.

**Figure S4:**
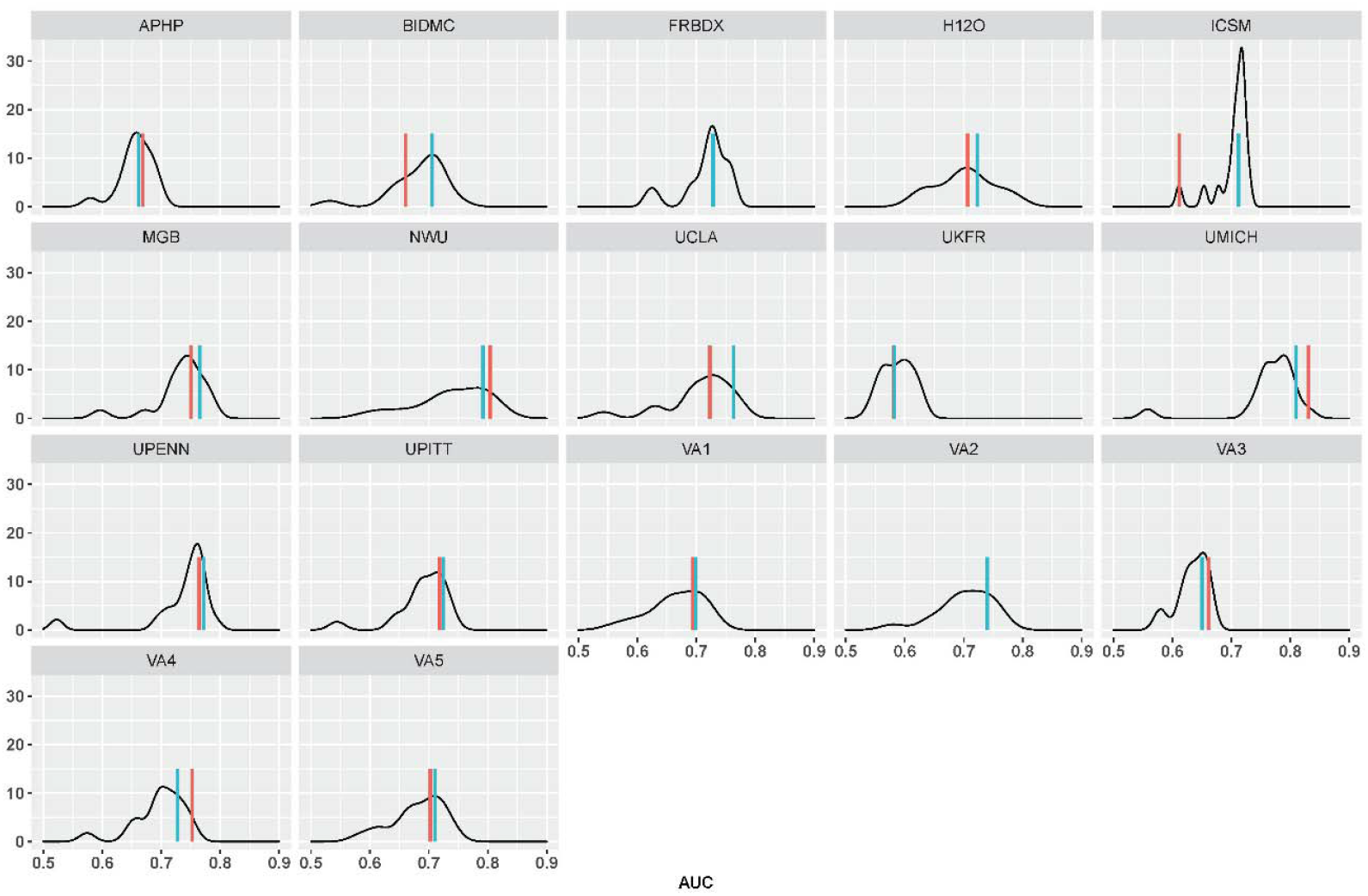
Comparing the AUC of locally trained model (red), the SurvMaximin model (blue), and the risk models trained in each external site (black) for predicting 14-day mortality by using a given health care center (APHP, FRBDX, UKFR, BIDMC, MGB, UPENN, UPITT, NWU, UMICH, UCLA, VA1 - VA5) as a potential target site.

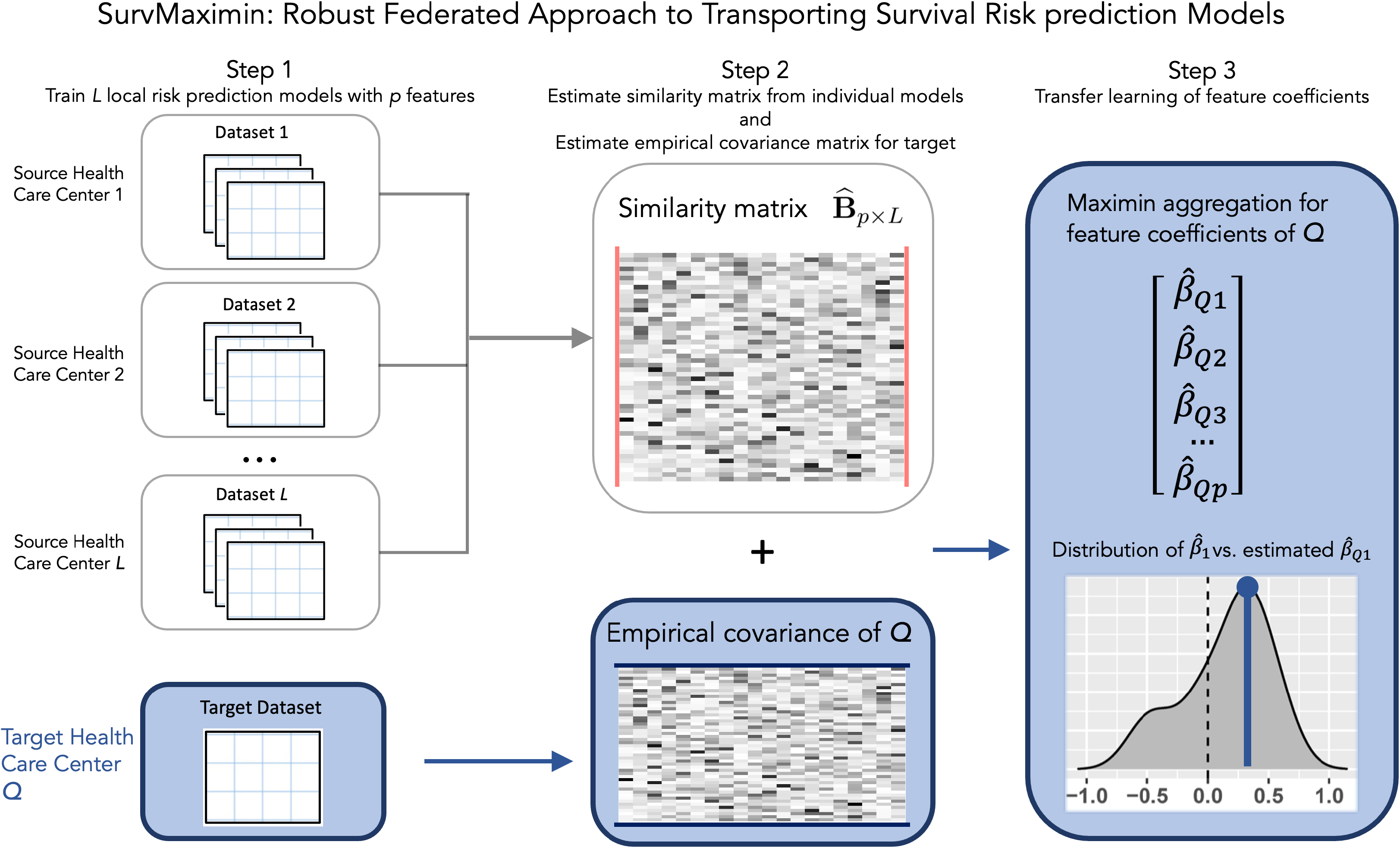

## Notes

### Competing Interest Statement

The authors have declared no competing interest.

### Author Declarations

IRB Approval was obtained at Assistance Publique - Hopitaux de Paris, Beth Israel Deaconess Medical Center, Bordeaux University Hospital, Instituti Instituti Clinici Scientifici Maugeri Hospitals, University of Kansas Medical Center, Massachusetts General Brigham, Northwestern University, Medical Center University of Freiburg, University of Pittsburgh, and VA North Atlantic, Southwest, Midwest, Continental and Pacific. An exempt determination was made by Institutional Review Boards at Hospital Universitario 12 de Octubre, University of California Los Angeles, University of Michigan, and University of Pennsylvania.

## References

1. David R Cox. “Regression models and life-tables”. In: Journal of the Royal Statistical Society: Series B (Methodological) 34.2 (1972), pp. 187–202.

2. Richard A Deyo, Daniel C Cherkin, and Marcia A Ciol. “Adapting a clinical comorbidity index for use with ICD-9-CM administrative databases”. In: Journal of clinical epidemiology 45.6 (1992), pp. 613–619.

3. Hal Daumé III. “Frustratingly easy domain adaptation”. In: arXiv preprint 0907.1815 (2009).

4. Shawn N Murphy et al. “Serving the enterprise and beyond with informatics for integrating biology and the bedside (i2b2)”. In: Journal of the American Medical Informatics Association 17.2 (2010), pp. 124–130.

5. Phyllis Torda, Esther S Han, and Sarah Hudson Scholle. “Easing the adoption and use of electronic health records in small practices”. In: Health Affairs 29.4 (2010), pp. 668–675.

6. Michael Wolfson et al. “DataSHIELD: resolving a conflict in contemporary bioscience?performing a pooled analysis of individual-level data without sharing the data”. In: International journal of epidemiology 39.5 (2010), pp. 1372–1382.

7. Hajime Uno et al. “On the C-statistics for evaluating overall adequacy of risk prediction procedures with censored survival data”. In: Statistics in medicine 30.10 (2011), pp. 1105–1117.

8. Stef Van Buuren and Karin Groothuis-Oudshoorn. “mice: Multivariate imputation by chained equations in R”. In: Journal of statistical software 45.1 (2011), pp. 1–67.

9. Sandra L Decker, Eric W Jamoom, and Jane E Sisk. “Physicians in nonprimary care and small practices and those age 55 and older lag in adopting electronic health record systems”. In: Health Affairs 31.5 (2012), pp. 1108–1114.

10. Yuan Wu et al. “G rid Binary LO gistic RE gression (GLORE): building shared models without sharing data”. In: Journal of the American Medical Informatics Association 19.5 (2012), pp. 758–764.

11. Peter Bühlmann and Nicolai Meinshausen. “Magging: maximin aggregation for inhomogeneous large-scale data”. In: arXiv preprint 1409.2638 (2014).

12. Yolanda Hagar et al. “Survival analysis with electronic health record data: Experiments with chronic kidney disease”. In: Statistical Analysis and Data Mining: The ASA Data Science Journal 7.5 (2014), pp. 385–403.

13. Chia-Lun Lu et al. “WebDISCO: a web service for distributed cox model learning without patient-level data sharing”. In: Journal of the American Medical Informatics Association 22.6 (2015), pp. 1212–1219.

14. Nicolai Meinshausen and Peter Bühlmann. “Maximin effects in inhomogeneous large-scale data”. In: The Annals of Statistics 43.4 (2015), pp. 1801–1830.

15. Dominik Rothenhäusler, Nicolai Meinshausen, and Peter Bühlmann. “Confidence intervals for maximin effects in inhomogeneous large-scale data”. In: Statistical Analysis for High-Dimensional Data. Springer, 2016, pp. 255–277.

16. Yan V Sun and Yi-Juan Hu. “Integrative analysis of multi-omics data for discovery and functional studies of complex human diseases”. In: Advances in genetics 93 (2016), pp. 147–190.

17. Young-Gun Kim et al. “Rate of electronic health record adoption in South Korea: a nation-wide survey”. In: International journal of medical informatics 101 (2017), pp. 100–107.

18. Jorge Tavares and Tiago Oliveira. “Electronic health record portal adoption: a cross country analysis”. In: BMC medical informatics and decision making 17.1 (2017), pp. 1–17.

19. Turki Turki, Zhi Wei, and Jason TL Wang. “Transfer learning approaches to improve drug sensitivity prediction in multiple myeloma patients”. In: IEEE Access 5 (2017), pp. 7381–7393.

20. Weihua Hu et al. “Does distributionally robust supervised learning give robust classifiers?” In: International Conference on Machine Learning. PMLR. 2018, pp. 2029–2037.

21. Chengchun Shi et al. “Maximin projection learning for optimal treatment decision with heterogeneous individualized treatment effects”. In: Journal of the Royal Statistical Society. Series B, Statistical methodology 80.4 (2018), p. 681.

22. Trevor Hastie, Robert Tibshirani, and Martin Wainwright. Statistical learning with sparsity: the lasso and generalizations. Chapman and Hall/CRC, 2019.

23. Shiori Sagawa et al. “Distributionally robust neural networks for group shifts: On the importance of regularization for worst-case generalization”. In: arXiv preprint 1911.08731 (2019).

24. Gaurav Singal et al. “Association of patient characteristics and tumor genomics with clinical outcomes among patients with non–small cell lung cancer using a clinicogenomic database”. In: Jama 321.14 (2019), pp. 1391–1399.

25. Gabriel A Brat et al. “International electronic health record-derived COVID-19 clinical course profiles: the 4CE consortium”. In: Npj Digital Medicine 3.1 (2020), pp. 1–9.

26. Rui Duan et al. “Learning from local to global: An efficient distributed algorithm for modeling time-to-event data”. In: Journal of the American Medical Informatics Association 27.7 (2020), pp. 1028–1036.

27. Zijian Guo. “Inference for High-dimensional Maximin Effects in Heterogeneous Regression Models Using a Sampling Approach”. In: arXiv preprint 2011.07568 (2020).

28. Ilker Kose et al. “Adoption rates of electronic health records in Turkish Hospitals and the relation with hospital sizes”. In: BMC Health Services Research 20.1 (2020), pp. 1–16.

29. Sai Li, T Tony Cai, and Hongzhe Li. “Transfer learning for high-dimensional linear regression: Prediction, estimation, and minimax optimality”. In: arXiv preprint 2006.10593 (2020).

30. Hamsa Bastani. “Predicting with proxies: Transfer learning in high dimension”. In: Management Science 67.5 (2021), pp. 2964–2984.

31. T Tony Cai and Hongji Wei. “Transfer learning for nonparametric classification: Minimax rate and adaptive classifier”. In: The Annals of Statistics 49.1 (2021), pp. 100–128.

32. Tianxi Cai, Molei Liu, and Yin Xia. “Individual data protected integrative regression analysis of high-dimensional heterogeneous data”. In: Journal of the American Statistical Association (2021), pp. 1–1

33. Griffin M Weber et al. “International Changes in COVID-19 Clinical Trajectories Across 315 Hospitals and 6 Countries: a 4CE Consortium Study.” In: J. med. internet res (2021).

